# The effectiveness of school-based obesity prevention interventions on psychosocial and educative outcomes in children aged 6-18 years: a secondary data analysis

**DOI:** 10.1101/2025.07.10.25331240

**Authors:** McDiarmid Katrina, Tara Clinton-McHarg, Kate M O’Brien, Kate Bartlem, Luke Wolfenden, Ashley Blowes, Caroline Kuhne, Daniel Chun-Wei Lee, Rebecca K Hodder

**Affiliations:** National Centre of Implementation Science: Booth Building, Wallsend Health Services, Longworth Avenue, Wallsend NSW 2287, Australia; School of Medicine and Public Health, College of Health Medicine and Wellbeing, University of Newcastle: University Dr, Callaghan NSW 2308, Australia; Melbourne School of Population and Global Health, Faculty of Medicine, Dentistry and Health Sciences, University of Melbourne: Parkville VIC 3052, Australia; School of Psychological Sciences, College of Engineering Science and Environment, University of Newcastle: University Dr, Callaghan NSW 2308, Australia; Hunter Medical Research Institute: Lot 1 Kookaburra Cct, New Lambton Heights NSW 2305, Australia

**Author notes:** Corresponding author: Katrina McDiarmid (; 0403963012).

**Keywords:** Obesity, child, school, education, mental health, wellbeing

## Abstract

**Background:** Childhood obesity remains a global public health priority. The two main risk behaviours for obesity, poor diet and physical inactivity, are both often targeted for obesity prevention. These risk behaviours may also influence other psychosocial and educative outcomes; however, few studies have examined the extent to which interventions targeting obesity also impact on these outcomes. This review aims to synthesise the effects of school-based child obesity prevention interventions on psychosocial and educative outcomes.

**Methods:** A secondary data analysis of an existing systematic review of childhood obesity prevention interventions was conducted. Additional literature searches identified studies published between 1990 and 2023 associated with the originally included studies. Studies were eligible if they reported any psychosocial or educative outcomes. Outcome data at follow-up were extracted. Where possible, meta-analysis was conducted to calculate standardised mean differences (SMD), otherwise narrative synthesis was conducted.

**Results:** Thirty-two studies were eligible for inclusion. The following psychosocial outcomes were meta-analysed: quality of life (SMD 0.05; 95% CI (0.01, 0.1)), wellbeing (SMD 0.14; 95% CI (−0.03, 0.3)), self-esteem (SMD 0.07; 95% CI (−0.06, 0.19)), self-worth (SMD 1.12; 95% CI (−3.41, 5.64)), and body satisfaction (SMD –0.06; 95% CI (−0.23, 0.11)). The following educative outcomes were meta-analysed: overall academic attainment (SMD 0.82; 95% CI (−1.02, 2.66)), maths (SMD 0; 95% CI (−0.02, 0.02)) and reading (SMD 0.1; 95% CI (−0.18, 0.39)).

**Conclusion:** School-based obesity prevention interventions may have small benefits for children’s psychosocial outcomes however findings for educative outcomes remain uncertain; further research is needed to clarify and strengthen this finding.

## Introduction

The high prevalence of obesity is increasing the global burden of chronic diseases such as diabetes, cancer and cardiovascular disease [1]. Obesity in childhood is of particular concern as children who are overweight or obese are likely to remain overweight or obese throughout their lifespan [2]. In 2020, 175 million children and adolescents worldwide were living with obesity [3]. As a result, international guidelines recommend obesity prevention interventions for children and adolescents to reduce the lifelong burden on both individuals and the community [4, 5].

Schools are a logical setting for delivering obesity prevention interventions, as they provide access to children and adolescents for prolonged periods each day [6]. These years are a critical development period where lifestyle habits are being established and as such, supporting student health and wellbeing is an important part of a school’s role [5, 7]. A recent systematic review found that school-based obesity prevention interventions overall have a small positive effect on body mass index (BMI) in comparison to control groups (standardised mean difference (SMD) –0.03, 95% CI –0.06 to –0.01; moderate certainty evidence) [8], supporting the use of schools as an intervention setting.

However, weight status is not the only consideration when measuring the impact of obesity on health and well-being. Children and adolescents who are overweight or obese are also more likely to experience mental health symptoms such as anxiety and depression, lower self-reported quality of life and lower self-esteem than those within a healthy weight range [9–14]. Educative outcomes such as school academic performance, are also influenced by obesity although the evidence is mixed. A review of cross-sectional and longitudinal studies found that obesity had a negative impact on academic achievement, as measured through grades, in 15 of 34 identified studies [15] Other reviews focussing on longitudinal studies also found a small impact of obesity on academic achievement [16–18] with two reviews establishing a more significant impact on girls than boys [17, 18].

The primary objective of obesity prevention interventions is usually to prevent excess weight gain, and interventions often focus on two key modifiable risk behaviours: poor diet (i.e. excessive energy intake and low fruit and vegetable consumption), and physical inactivity (i.e. low levels of activity and high levels of sedentary behaviour) [19, 20]. As such, diet and physical activity are frequently targeted in school-based interventions aiming to address and prevent obesity in children and adolescence. Schools are encouraged to address these behaviours as part of the World Health Organisation’s Health Promoting Schools Framework where health is promoted through the curriculum, school environment and broader school community [21].

However, there is growing evidence that improving dietary and physical activity outcomes may also improve psychosocial outcomes in children and adolescents [22–30]. For example, systematic review evidence in children aged 3-18 years reported that those consuming a healthy diet also had better quality of life than those not consuming a healthy diet [30] and those that reported higher levels of physical activity also reported higher quality of life scores [29]. Another systematic review assessing physical activity interventions in general school populations found small improvements in mental health symptomology and psychological wellbeing [25]. Given this association and that these two behaviours are typically targeted in obesity prevention interventions, obesity prevention interventions may also positively impact psychosocial outcomes in children and adolescence.

Healthy eating and physical activity also have an impact on educative outcomes for children and adolescents. Systematic reviews of intervention studies report evidence which suggests that increased physical activity has a positive impact on academic achievement [28, 31–33]. An umbrella review investigating the effect of school physical activity on academic achievement found consistent small to medium positive effects of increased physical education classes and active classrooms on both specific school subject performance and overall academic achievement [34]. Previous cross-sectional and longitudinal evidence also suggests that higher diet quality has a beneficial impact on academic performance [31, 35–39] however there have been limited trials testing the effect of a nutrition intervention, excluding the provision of nutritionally balanced school meals, on academic achievement. One systematic review assessing the impact of dietary interventions on academic achievement in overweight or obese children only, found the mean average in academic achievement was 0.32 standard deviations higher for those in the intervention group compared with the control [40]. Therefore, as with psychosocial outcomes, obesity prevention interventions targeting diet and physical activity may extend their benefits to educative outcomes as well.

Given the absence of up-to-date reviews examining the effects of obesity prevention interventions delivered in the school context on psychosocial and educative outcomes, the aims of this review were to conduct a secondary data analysis of school-based child obesity prevention interventions to determine their impact on a range of psychosocial and educative outcomes.

## Methods

A systematic review was previously conducted to assess the effectiveness of childhood obesity prevention interventions on child weight status (e.g. BMI) [8]. This current review reports a secondary data analysis of studies included in the original systematic review, to determine the additional impact that obesity interventions may have on student psychosocial and educative outcomes. The protocol for this secondary data analysis was prospectively registered (PROSPERO 2021 CRD42021281106) [41].

## Study inclusion criteria

Both the original systematic review and this review included randomised controlled trials (RCTs) that compared obesity prevention interventions targeting diet and/or physical activity behaviours, with a control group (no intervention or usual care or attention control). RCTs published before 1990 were excluded as per the exclusion criteria from the original review.

### Participants

The participants in included studies were children of any weight with a mean age between six and 18 years at baseline. Studies that only enrolled children who were overweight or obese at baseline were excluded, as these trials are considered to be focused on obesity treatment rather than obesity prevention. Studies which were designed to prevent obesity in pregnant women or designed for children with a critical illness or severe co-morbidities, were also excluded.

### Intervention

Consistent with the original review, eligible interventions in this study were those that:

1. Were designed or had an underlying intention to prevent obesity: this includes studies that targeted diet only, physical activity only, or both;
2. Had an active intervention period of any duration and reported follow-up outcome data at a minimum of 12 weeks from baseline;
3. Randomly assigned individuals or groups of individuals to an experimental group, however, for those with group randomisation, only cluster-RCTs (C-RCTs) with six or more groups were eligible; and
4. Were delivered by any personnel, for example, researchers, primary care physicians (general practitioners), nutrition/diet professionals, teachers, physical activity professionals, health promotion agencies, health departments, faith leaders or others.

In the original systematic review, interventions conducted in any setting were eligible for inclusion (e.g. schools, community, after-school). However, for this secondary data analysis only studies conducted in school settings were eligible, inclusive of kindergarten, primary, middle and secondary schools. These included interventions conducted both during school hours and out-of-school-hours care.

### Outcomes

Studies in the original review were required to report both baseline and post-intervention data for one or more of the following obesity related outcomes: BMI/zBMI score and/or prevalence of overweight and obesity.

To be eligible for this secondary data analysis, studies must have additionally reported the effect of an intervention on one or more of the following eligible psychosocial outcomes:

– wellbeing, as social and emotional wellbeing
– quality of life, as a culmination of a person’s mental and physical wellbeing
– mental health symptomology, including measures of anxiety and depression, and problem behaviours
– body satisfaction, as differences between desired body shape and perceived body shape
– self-worth, as global self-worth measured through perceptions of worthiness as a person
– self-esteem, as a sense of value about oneself

Or one or more of the following educative outcomes:

– school grades, including overall grades and individual subjects
– classroom behaviour, measures of disruptiveness
– cognitions and engagement, as ability to stay focused in the classroom.

## Search methods

The search strategy for the original review involved searches of electronic databases including the Cochrane Central Register of Controlled Trials (CENTRAL) in the Cochrane Library, Medline, Embase, Cumulative Index to Nursing and Allied Health Literature (CINAHL) and PsycINFO [8]. Studies were not excluded based on language.

For this secondary data analysis, additional database searches, including searches of ClinicalTrials.gov and the WHO International Clinical Trials Registry Platform for associated trial registries, were also conducted to identify any other potentially eligible associated records for each study that was included in the original review. Authors of all studies included in the original review were contacted via email to obtain any associated publications or relevant unpublished data prior to screening for eligible studies.

## Study selection

All included studies and their associated records from the original review, as well as any new records found from the additional searches, were independently screened by two review authors (KM and DCWL) using a standardised screening tool to assess eligibility. Any differences were resolved through discussion, and if required, consultation with a third review author (RH) to resolve any disagreements.

## Data collection

Three review authors (KM, KB and AB) independently extracted the outcome data from each included study using a standardised data extraction tool in REDCap [42]. Any differences in data extraction between review authors were resolved via discussion or with another reviewer (RH). Data regarding study characteristics (design, participants, interventions, weight outcomes, cost and adverse effects) were available from the original published review. Information relevant to assessing risk of bias of the psychosocial and educative outcomes were also extracted.

## Assessment of risk of bias and quality of evidence

Risk of bias of included studies was assessed using the first version of the Cochrane Handbook for Systematic Reviews of Interventions risk of bias tool, in line with the methods of the original review [43]. Risk of bias assessments reported in the original systematic review were adopted for this secondary data analysis for the following domains: sequence generation, allocation sequence concealment, blinding of participants and personnel, and other biases. Risk of bias was re-assessed for psychosocial and educative outcomes in this secondary data analysis for the following domains: blinding of outcome assessment, incomplete outcome data, and selective outcome reporting. Risk of bias was assessed independently by two review authors (KM, TCM) and summarised in tabular form. Any disagreements in risk of bias assessments were resolved via discussion, or consultation with a third reviewer (RH).

The Grading of Recommendations Assessment, Development and Evaluation (GRADE) approach was used to assess the quality of the evidence for each eligible psychosocial and educative outcome across the trials included in the secondary data analysis. The GRADE assessment was completed by two review authors (KM, RH) for each outcome with first preference for the meta-analysis results, then results from synthesis without meta-analysis (SWiM), then for individual studies. Any disagreements in GRADE assessments were resolved by discussion. The certainty of the body of evidence for each outcome was graded as ‘high’, ‘moderate’, ‘low’, or, ‘very low’.

## Data analysis and synthesis

Each outcome was synthesised by comparing the intervention groups to the control groups for all intervention types. For studies with multiple intervention arms or subgroup results, data were combined using formulas provided in the Cochrane Handbook [44]. Where multiple measures for a psychosocial outcome were reported within a study, self-report measures were chosen over proxy-measure. Where multiple measures for an educative outcome were reported within a study, objective measures were chosen over subjective measures. Where there were multiple measures of the same outcome (at the same level of the decision hierarchy), one outcome measure was randomly selected via a random number generator. Outcomes were selected for each study by two authors (KM, KO) following this decision hierarchy. For each study reporting on multiple time points, data was extracted for the first follow up only, as per the methods of the original review.

Outcomes that were reported in three or more studies were eligible for meta-analysis. Analysis was conducted using R version 4.4.1 (2024-06-14). SMDs were used to compare various outcome measures, enabling different types of outcomes measures to be analysed on a common scale. For continuous data, the approach to calculating SMDs varied depending on the available data. Where SMDs were reported in the original studies, these were extracted directly. Where not reported, SMDs and their sampling variances were calculated using the *escalc* function from the *metafor* package in R [45]. When necessary, standard deviations were derived from confidence intervals or standard errors using methods described in the Cochrane Handbook [46]. For categorical outcomes, SMDs or odds ratios (ORs) were extracted when provided. When only raw group data were available, ORs were calculated directly, and SMDs were derived where appropriate.

C-RCTs were checked if they appropriately adjusted for clustering. If not, clustering was accounted for by inflating the standard errors using the design effect, calculated as: *DE* = 1 + (*m* − 1) × *ICC*, where *m* was the average cluster size and *ICC* was the intraclass correlation coefficient [44]. When *ICC* values were not reported, estimated values were obtained from studies with similar designs.

Random effects meta-analyses were used to pool the standardized mean differences (SMDs) and their variances for all outcomes using the R package ‘metafor’ [45]. The random effects model was chosen since the included studies varied in their populations, intervention types, and settings, making it reasonable to assume that the true effect sizes differ between studies.

Results are presented as a pooled SMD with a 95% confidence interval. A SMD of 0.2 is considered small, 0.5 is considered a moderate effect and 0.8 a large effect [47]. To quantify heterogeneity, *I*^2^ (the percentage of total variation across studies due to heterogeneity) is reported with the respective confidence intervals. Subgroup analysis between intervention target (diet, physical activity, or both) was performed to explore the reasons for high heterogeneity (*I*^2^ ≥ 50%).

## Results

### Study characteristics

A total of n=824 unique records, including n=478 studies from the original review as well as n=346 potentially associated records identified through the updated literature search and author contact, were screened against the eligibility criteria. After title and abstract screening, n=460 studies were included for full text screening against the eligibility criteria. Of these, n=428 individual studies were further excluded. Thirty-two studies (from n=94 records) were deemed eligible for inclusion.

Of the 32 studies, five were RCTs and 27 were C-RCTs that compared one or more active intervention groups to a control. A total of 19,304 participants with an average age of 10.4 years and 66 trial arms were analysed as part of the 32 studies. All studies were conducted in schools whether during school time (n=27) or as an after-school program (n=5). Twenty-one were conducted in a primary school setting, and 11 were conducted in a secondary school setting. Most studies were from high income countries (United States of America (n=13), Australia (n=7), United Kingdom (n=4), Denmark (n=1), England (n=1), Netherlands (n=1), Switzerland (n=1)), with the remainder from upper middle income countries (Brazil (n=2), China (n=1), South Africa (n=1)). One study targeted diet only, 14 targeted physical activity only and 17 studies targeted both diet and physical activity. The duration of these interventions ranged from 5 weeks to 3.5 years. Twenty-five studies reported a psychosocial outcome, and 10 reported an educative outcome (Appendix I, Table S1).

**Fig 1.**
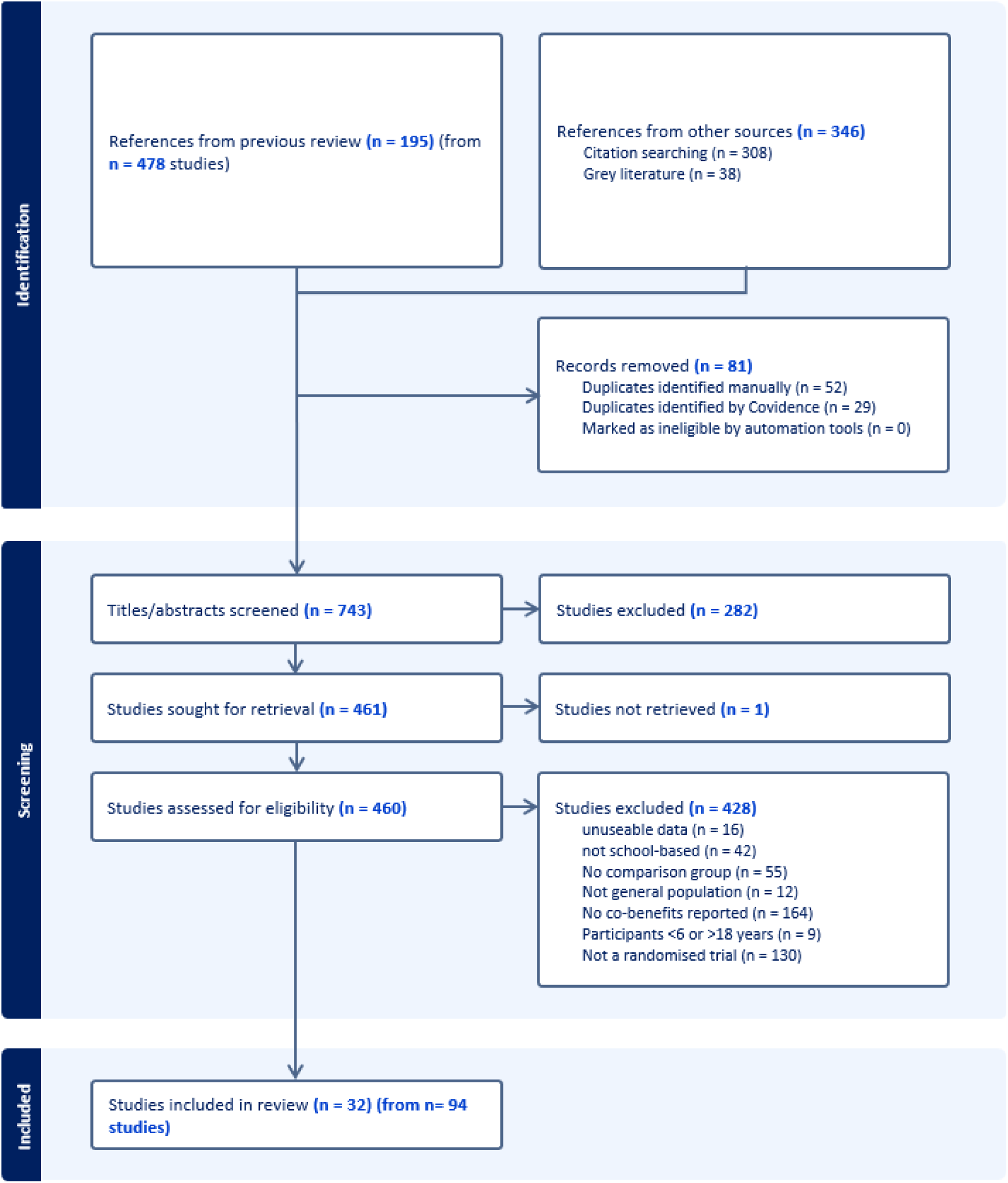
Study flow-chart – flow of studies through identification and screening.

### Risk of bias

Overall risk of bias was deemed high for quality of life (n=10/10 studies), wellbeing (n=5/5 studies), self-esteem (n=5/5 studies), body satisfaction (n=9/9 studies), engagement (n=1/1 study), problem behaviours (n=1/1 study), overall academic attainment (n=4/4 studies), and attention and behaviour (n=2/2 studies). Overall risk of bias was deemed unclear for anxiety (n=1 study). Overall risk of bias was mixed for self-worth (n=4 high, n=1 unclear), depression (n=1 high, n=1 unclear), maths (n=4 high, n=3 unclear), reading (n=4 high, n=3 unclear), and writing (n=2 high, n=2 unclear) (Appendix II, Table S2).

### Psychosocial outcomes

#### Quality of life

Ten studies assessed the effect of an obesity prevention intervention on quality of life. There was a small but statistically significant increase in quality of life (SMD 0.05; 95% CI (0.01, 0.1); n=9; participants=8210; p-value 0.02; low certainty evidence) (Table 1). Heterogeneity was low (I^2^ 0; 95% CI (0, 85.5)).

**Table 1.** Overall meta-analysis results for psychosocial and educative outcomes of randomised controlled obesity prevention interventions published from 1990-2023.

| Outcome | Studies | Participants | SMD (95% CI) | I<sup>2</sup> (95% CI) | GRADE and direction of effect |
| --- | --- | --- | --- | --- | --- |
| Quality of life | 9 | 8210 | 0.05 (0.01, 0.1)* | 0 (0, 85.5) | Low<br>▲ |
| Wellbeing | 5 | 4955 | 0.14 (-0.03, 0.3) | 61.7 (0, 95.6) | Very low<br>▲ |
| Self-esteem | 5 | 2539 | 0.07 (-0.06, 0.19) | 35.2 (0, 91.8) | Very low<br>▲ |
| Self-worth | 3 | 1519 | 1.12 (-3.41, 5.64) | 98.2 (93.4, 99.9) | Very low<br>▲ |
| Body satisfaction | 7 | 3531 | -0.06 (-0.23, 0.11) | 76.1 (42.4, 98.4) | Very low<br>▲ |
| Overall academic attainment | 4 | 3704 | 0.82 (-1.02, 2.66) | 98.9 (96.6, 99.9) | Very low<br>▲ |
| Maths | 5 | 2256 | 0 (-0.02, 0.02) | 0 (0, 0) | Very low<br>◀▶ |
| Reading | 4 | 1557 | 0.1 (-0.18, 0.39) | 0 (0, 86.7) | Very low<br>▲ |

SMD = standardised mean difference; \* denotes statistically significant finding; ▲ = positive direction of effect, ▼ = negative direction of effect, ▶▶ = no change/mixed effects/conflicting findings

One study could not be included in meta-analysis (authors only reported a direction of effect) [48]. This study favoured the intervention compared to the control (Table 2).

**Table 2.** Synthesis without meta-analysis (direction of effect plot of intervention group) of select psychosocial and educative outcomes from randomised controlled obesity interventions published since 1990.

| Study identification | Study design | No. participants: Intervention/control | Quality of life | Problem behaviours | Engagement |
| --- | --- | --- | --- | --- | --- |
| Clemes 2020 | C-RCT | 84/86 | ▲ | ▼ | ◀▶ |

C-RCT = cluster-randomised controlled trial; ▲ = positive direction of effect, ▼ = negative direction of effect, ▶▶ = no change/mixed effects/conflicting findings

#### Wellbeing

Five studies assessed the effect of an obesity prevention intervention on wellbeing. There was an increase in wellbeing across all studies, but this was not statistically significant (SMD 0.14; 95% CI (−0.03, 0.3); n=5; participants=4955; p-value 0.12; very low certainty evidence) (Table 1). Subgroup analysis was unable to be completed to explore high heterogeneity due to the limited number of included studies reporting on wellbeing (I^2^ 61.7; 95% CI (0, 95.6)).

#### Self-esteem

Five studies assessed the effect of an obesity prevention intervention on self-esteem (Table 1). There was a small increase in self-esteem across all studies, but this was not statistically significant (SMD 0.07; 95% CI (−0.06, 0.19); n=5; participants=2539; p-value 0.31; very low certainty evidence). Heterogeneity was low (I^2^ 35.2; 95% CI (0, 91.8)).

#### Self-worth

Five studies assessed the effect of an obesity prevention intervention on self-worth (Table 1). There was a large increase in self-worth but this was not statistically significant (SMD 1.12; 95% CI (−3.41, 5.64); n=3; participants=1519; p-value 0.4; very low certainty evidence). Subgroup analysis was unable to be completed to explore high heterogeneity due to the limited number of included studies reporting on self-worth (I^2^ 98.2; 95% CI (93.4, 99.9)).

Two studies could not be included in meta-analysis due to limited data [49, 50]. The combination of p-values suggests that there is no evidence of benefit of obesity prevention interventions on self-worth (p=0.05; n=2; participants=535).

#### Body satisfaction

Nine studies assessed the effect of an obesity prevention intervention on body satisfaction (Table 1). There was a small decrease in body satisfaction scores, but this was not statistically significant (SMD –0.06; 95% CI (−0.23, 0.11); n=7; participants=3531; p-value 0.48; very low certainty evidence). Subgroup analysis was unable to be completed to explore high heterogeneity due to the limited number of included studies reporting on body satisfaction (I^2^ 76.1; 95% CI (42.4, 98.4)).

Two studies could not be included in meta-analysis due to limited data [49, 50]. The combination of p-values suggests that there is no evidence of benefit of obesity prevention interventions on body satisfaction (p=0.14; n=2; participants=535).

#### Anxiety

One study assessed the effect of an obesity prevention intervention on anxiety [51]. Authors assessed the impact of an obesity prevention intervention (Creating Opportunities for Personal Empowerment) compared to an attention control group (Healthy Teens program) and reported no difference between groups at follow-up (mean difference (MD) 0.46; 95% CI –0.79, 1.7; moderate certainty evidence).

#### Depression

Two studies assessed the effect of an obesity prevention intervention on depression [51, 52]. Authors provided sufficient data to summarise effect estimates. The SMD effect sizes ranged from 0.03 (95% CI –0.09, 0.14) to 0.07 (95% CI –0.09, 0.23) (n=2, participants=1941; very low certainty evidence).

#### Problem behaviours

One study assessed the effect of an obesity prevention intervention on problem behaviours and only reported a direction of effect [48]. This study favoured the intervention compared to the control (low certainty evidence) (Table 2).

## Educative outcomes

### Overall academic attainment

Four studies assessed the effect of obesity prevention interventions on overall academic attainment (Table 1). There was a positive impact on academic scores across all studies, but this was not statistically significant (SMD 0.82; 95%CI (−1.02, 2.66); n=4; participants=3704; p-value 0.25; very low certainty evidence). Subgroup analysis was unable to be completed to explore high heterogeneity due to the limited number of included studies reporting on overall academic attainment (I^2^ 98.9; 95% CI (96.6, 99.9)).

### Maths

Seven studies assessed the effect of obesity prevention interventions on maths scores (Table 1). There was no change in maths scores (SMD 0; 95% CI (−0.02, 0.02); n=5; participants=2256; p-value 0.86; very low certainty evidence). Heterogeneity was low (I^2^ 0; 95% CI (0, 0)).

Two studies could not be included in meta-analysis due to limited data [53, 54]. The combination of p-values suggests that there is strong evidence of benefit of obesity prevention interventions on maths scores in at least one study (p<0.01, n=2, participants=823).

### Reading

Six studies assessed the effect of obesity prevention interventions on reading scores (Table 1). There was a small increase in reading scores, but this was not statistically significant (SMD 0.1; 95% CI (−0.18, 0.39); n=4; participants=1557; p-value 0.33; very low certainty evidence). Heterogeneity was low (I^2^ 0; 95% CI (0, 96.7)).

Two studies could not be included in meta-analysis due to limited data [53, 54]. The combination of p-values suggests that there is strong evidence of benefit of obesity prevention interventions on reading scores in at least one study (p=0.03, n=2, participants=823).

### Writing

Four studies assessed the effect of obesity prevention interventions on writing scores but could not be included in meta-analysis due to limited data [48, 53–55]. The combination of p-values suggests that there is strong evidence of benefit of obesity prevention interventions on writing scores in at least one study (p=0.04, n=4, participants=1413; low certainty evidence).

### Attention and behaviour

Two studies assessed the effect of obesity prevention interventions on attention and behaviour [56, 57]. Authors provided enough data to summarise effect estimates. The beta effect sizes ranged from –0.16 (95% CI –1.29, 0.98) to 2.93 (95% CI –5.01, 10.86) (n=2, participants= 1486; very low certainty evidence).

### Engagement

One study assessed the effect of an obesity prevention intervention on engagement. Authors assessed the impact of an obesity prevention intervention compared to usual care control and reported no difference in change from baseline between groups (MD 0; 95% CI –0.17, 0.17; low certainty evidence) (Table 2).

## Discussion

This review examined the effects of school-based obesity prevention interventions on a range of psychosocial and educative outcomes. The review found that these interventions may lead to slight reductions in anxiety and problem behaviours, as well as slight improvements in quality of life and writing. These interventions may also lead to a slight increase in wellbeing, self-esteem, self-worth, body satisfaction, overall academic attainment, and reading, however this evidence is uncertain. There was no evidence of a beneficial effect in maths scores or classroom engagement, and findings were mixed for attention and behaviour. Further, a small, uncertain negative effect on depression was observed. However, most of these findings were based on low to very low certainty evidence.

Psychosocial outcomes have previously been assessed in a 2009 review of school-based obesity prevention interventions in children and adolescents. The review did not find any significant effects on psychosocial outcomes including body image, self-worth or self-esteem and did not quantify the direction of effect for these outcomes [58]. In contrast, the present study did identify small positive effects for several psychosocial outcomes, although most effect sizes were small and statistically non-significant.

Outside of an obesity context, previous reviews have examined the effectiveness of school-based physical activity interventions on psychosocial outcomes. A 2020 review found beneficial effects on anxiety and wellbeing, but no evidence of effects on depression, quality of life, self-esteem or self-worth [59]. The interventions included were described as improving physical strength and the certainty of the evidence was not assessed. Another review of physical activity interventions focussing on varied exercise modalities and intensities found significant positive effects on anxiety, depression, and self-esteem [60]. However, most included studies were not school-based, and the overall certainty of the evidence was low. No reviews were found that assessed the effects of school-based nutrition interventions on psychosocial outcomes.

To our knowledge, this is the first review to synthesise the effects of school-based obesity prevention interventions on educative outcomes in a general population of children and adolescents. Several other reviews have assessed the effectiveness of school-based physical activity interventions separately and their effects on educative outcomes, but findings across these reviews have been inconsistent [61–63]. Similar to the findings of the current review, previous reviews have been limited by the variability in how academic achievement is measured. No reviews were found to assess the effect of school-based nutrition interventions on any educative outcomes.

A major limitation of the existing evidence base is the low number of studies reporting on psychosocial or educative outcomes following school-based obesity prevention interventions. This limited the scope of meta-analysis and sub-group analyses could not be performed, making it unclear whether targeting diet, physical activity, or both is more effective. Despite the known associations between these behaviours and psychosocial and educative outcomes, only 34% of school-based studies in the original review assessed psychosocial or academic outcomes. For schools following the Health Promoting Schools Framework, improved psychosocial and educative outcomes for school students are expected as a result [21].

Most of the findings of this review were assessed as low or very low certainty according to the GRADE approach, and most effect sizes were small and non-significant. Future high-quality trials in obesity prevention are encouraged to assess a broader range of psychosocial and educative outcomes to strengthen the evidence in this area.

The generalisability of these results is also limited. Most included studies were from high income countries with a primary school cohort aged 5-12 years. As such the findings may not be relevant to other populations, regions or socioeconomic contexts. Additionally, all included studies were conducted in the context of obesity prevention and reported assessing BMI only. Interventions that did not assess BMI, including general diet or physical activity interventions, were excluded from this review. This highlights an important gap in the literature and a future research opportunity to expand these findings.

### Conclusion

School-based obesity prevention interventions may have a small positive effect on selected psychosocial outcomes, including wellbeing, quality of life, self-esteem, self-worth, anxiety, problem behaviours and body dissatisfaction, and educative outcomes, including overall academic attainment, reading, and writing scores. Future research in obesity prevention should continue to explore the wider impacts of these interventions to better understand and strengthen the link between these health behaviours and broader psychosocial and educative outcomes. Stronger evidence in this area may further support the value of school-based obesity prevention initiatives.

## Statement regarding informed consent

For this type of study, formal consent is not required.

## Statement regarding ethical approval

This article does not contain any studies with human participants or animals performed by any of the authors.

## Author contribution statement

KM contributed to the conceptualisation and methods; conducted searches, screening of records, data extraction, interpretation of findings, risk of bias assessment, GRADE assessment; and led drafting of the manuscript. TCM contributed to the conceptualisation and methods, data extraction, interpretation of findings, and risk of bias assessment. KMO contributed to the conceptualisation and methods, data extraction, synthesis and interpretation of findings. KB contributed to the conceptualisation, and data extraction. LW contributed to the conceptualisation, and interpretation of findings. AB contributed to the conduct of searches, and data extraction. CK contributed to the data analysis and interpretation. DCWL contributed to the screening of records. RKH led the conceptualisation and supervision of lead author, and contributed to data extraction, interpretation of study findings, and GRADE assessment. All authors critically reviewed the manuscript.

## Data Availability

All data produced in the present study are available upon reasonable request to the authors

## Acknowledgements

KM is supported by a University of Newcastle RTP stipend. KB is supported by a NHMRC early career fellowship (APP1142272)

## References

1. World Health Organisation. Obesity and Overweight. World Health Organisation. 2021. https://www.who.int/news-room/fact-sheets/detail/obesity-and-overweight.

2. Lister NB, Baur LA, Felix JF, et al. Child and adolescent obesity. Nat Rev Dis Primers. 2023;9(1):24. doi:10.1038/s41572-023-00435-4

3. Lobstein T, Jackson-Leach R, Powis J, Brinsden H, Gray M. World obesity atlas 2023. 2023.

4. Stylianou M, Walker JL. An assessment of Australian school physical activity and nutrition policies. Australian and New Zealand journal of public health. 2018;42(1):16–21.

5. Langford R, Bonell C, Jones H, et al. The World Health Organization’s Health Promoting Schools framework: a Cochrane systematic review and meta-analysis. BMC public health. 2015;15(1):1–15.

6. Smith JD, Fu E, Kobayashi MA. Prevention and Management of Childhood Obesity and Its Psychological and Health Comorbidities. Annu Rev Clin Psychol. 2020;16:351–78. doi:10.1146/annurev-clinpsy-100219-060201

7. Pulimeno M, Piscitelli P, Colazzo S, Colao A, Miani A. School as ideal setting to promote health and wellbeing among young people. Health Promot Perspect. 2020;10(4):316–24. doi:10.34172/hpp.2020.50

8. Hodder RK, O’Brien KM, Lorien S, et al. Interventions to prevent obesity in school-aged children 6-18 years: An update of a Cochrane systematic review and meta-analysis including studies from 2015-2021. EClinicalMedicine. 2022;54:101635. doi:10.1016/j.eclinm.2022.101635

9. Chen Y, Zhang J, Yuan L, et al. Obesity and risk of depressive disorder in children and adolescents: A meta-analysis of observational studies. Child Care Health Dev. 2024;50(2):e13237. doi:10.1111/cch.13237

10. Ciezki S, Odyjewska E, Bossowski A, Glowinska-Olszewska B. Not Only Metabolic Complications of Childhood Obesity. Nutrients. 2024;16(4). doi:10.3390/nu16040539

11. Förster L-J, Vogel M, Stein R, et al. Mental health in children and adolescents with overweight or obesity. BMC Public Health. 2023;23(1). doi:10.1186/s12889-023-15032-z

12. Godina-Flores NL, Gutierrez-Gomez YY, Garcia-Botello M, Lopez-Cruz L, Moreno-Garcia CF, Aceves-Martins M. Obesity and its association with mental health among Mexican children and adolescents: systematic review. Nutr Rev. 2023;81(6):658–69. doi:10.1093/nutrit/nuac083

13. Oz B, Kivrak AC. Evaluation of depression, anxiety symptoms, emotion regulation difficulties, and self-esteem in children and adolescents with obesity. Arch Pediatr. 2023;30(4):226–31. doi:10.1016/j.arcped.2023.02.003

14. Rao WW, Zong QQ, Zhang JW, et al. Obesity increases the risk of depression in children and adolescents: Results from a systematic review and meta-analysis. J Affect Disord. 2020;267:78–85. doi:10.1016/j.jad.2020.01.154

15. Santana CCA, Hill JO, Azevedo LB, Gunnarsdottir T, Prado WL. The association between obesity and academic performance in youth: a systematic review. Obes Rev. 2017;18(10):1191–9. doi:10.1111/obr.12582

16. Elish P, Boedeker P, Lash TL, Gazmararian J. Longitudinal weight status and academic achievement in elementary schoolchildren in the United States. Int J Obes (Lond). 2023;47(7):644–50. doi:10.1038/s41366-023-01309-1

17. Martin A, Booth JN, McGeown S, et al. Longitudinal Associations Between Childhood Obesity and Academic Achievement: Systematic Review with Focus Group Data. Curr Obes Rep. 2017;6(3):297–313. doi:10.1007/s13679-017-0272-9

18. Watson A, D’Souza NJ, Timperio A, Cliff DP, Okely AD, Hesketh KD. Longitudinal associations between weight status and academic achievement in primary school children. Pediatr Obes. 2023;18(1):e12975. doi:10.1111/ijpo.12975

19. Australian Institute of Health and Wellbeing. Australian Burden of Disease Study 2018: key findings. Canberra 2021.

20. World health Organisation. Global status report on noncommunicable diseases 2014. Switzerland 2014.

21. Langford R, Bonell CP, Jones HE, et al. The WHO Health Promoting School framework for improving the health and well-being of students and their academic achievement. Cochrane Database Syst Rev. 2014;2014(4):Cd008958. doi:10.1002/14651858.CD008958.pub2

22. Chen L, Liu Q, Xu F, et al. Effect of physical activity on anxiety, depression and obesity index in children and adolescents with obesity: A meta-analysis. J Affect Disord. 2024;354:275–85. doi:10.1016/j.jad.2024.02.092

23. Liu L, Guo C, Lang F, Yan Y. Association of breakfast, total diet quality, and mental health in adolescents: a cross-sectional study of HBSC in Greece. Eur J Pediatr. 2023;182(12):5385–97. doi:10.1007/s00431-023-05180-0

24. Marsigliante S, Gomez-Lopez M, Muscella A. Effects on Children’s Physical and Mental Well-Being of a Physical-Activity-Based School Intervention Program: A Randomized Study. Int J Environ Res Public Health. 2023;20(3). doi:10.3390/ijerph20031927

25. Purgato M, Cadorin C, Prina E, et al. Umbrella Systematic Review and Meta-Analysis: Physical Activity as an Effective Therapeutic Strategy for Improving Psychosocial Outcomes in Children and Adolescents. J Am Acad Child Adolesc Psychiatry. 2024;63(2):172–83. doi:10.1016/j.jaac.2023.04.017

26. Rodrigues D, Machado-Rodrigues AM, Gama A, Silva MG, Nogueira H, Padez C. Body size, form, composition, and a healthy lifestyle associates with health-related quality of life among Portuguese children. Am J Hum Biol. 2023;35(8):e23902. doi:10.1002/ajhb.23902

27. Rodriguez-Ayllon M, Cadenas-Sanchez C, Estevez-Lopez F, et al. Role of Physical Activity and Sedentary Behavior in the Mental Health of Preschoolers, Children and Adolescents: A Systematic Review and Meta-Analysis. Sports Med. 2019;49(9):1383–410. doi:10.1007/s40279-019-01099-5

28. Spruit A, Assink M, van Vugt E, van der Put C, Stams GJ. The effects of physical activity interventions on psychosocial outcomes in adolescents: A meta-analytic review. Clin Psychol Rev. 2016;45:56–71. doi:10.1016/j.cpr.2016.03.006

29. Wu XY, Han LH, Zhang JH, Luo S, Hu JW, Sun K. The influence of physical activity, sedentary behavior on health-related quality of life among the general population of children and adolescents: A systematic review. PLoS One. 2017;12(11):e0187668. doi:10.1371/journal.pone.0187668

30. Wu XY, Zhuang LH, Li W, et al. The influence of diet quality and dietary behavior on health-related quality of life in the general population of children and adolescents: a systematic review and meta-analysis. Qual Life Res. 2019;28(8):1989–2015. doi:10.1007/s11136-019-02162-4

31. Bouchefra S, El Ghouddany S, Ouali K, Bour A. Is good dietary diversity a predictor of academic success? Acta Biomed. 2023;94(2):e2023014. doi:10.23750/abm.v94i2.13940

32. Enríquez-Del Castillo LA, Villegas-Balderrama CV, López-Alonzo SJ, Flores Olivares LA, Martínez-Trevizo A, Islas-Guerra SA. Planetary health diet versus usual diet in adolescents. How do food and physical activity influence academic performance? Nutr Hosp. 2024;41(1):28–37. doi:10.20960/nh.04614

33. Martin-Martinez C, Valenzuela PL, Martinez-Zamora M, Martinez-de-Quel O. School-based physical activity interventions and language skills: a systematic review and meta-analysis of randomized controlled trials. J Sci Med Sport. 2023;26(2):140–8. doi:10.1016/j.jsams.2022.12.007

34. Barbosa A, Whiting S, Simmonds P, Scotini Moreno R, Mendes R, Breda J. Physical Activity and Academic Achievement: An Umbrella Review. Int J Environ Res Public Health. 2020;17(16). doi:10.3390/ijerph17165972

35. Hayek J, de Vries H, Tueni M, Lahoud N, Winkens B, Schneider F. Increased Adherence to the Mediterranean Diet and Higher Efficacy Beliefs Are Associated with Better Academic Achievement: A Longitudinal Study of High School Adolescents in Lebanon. Int J Environ Res Public Health. 2021;18(13). doi:10.3390/ijerph18136928

36. Sánchez-Hernando B, Antón-Solanas I, Juárez-Vela R, et al. Healthy Lifestyle and Academic Performance in Middle School Students from the Region of Aragón (Spain). Int J Environ Res Public Health. 2021;18(16). doi:10.3390/ijerph18168624

37. Burrows T, Goldman S, Pursey K, Lim R. Is there an association between dietary intake and academic achievement: a systematic review. J Hum Nutr Diet. 2017;30(2):117–40. doi:10.1111/jhn.12407

38. MacLellan D, Taylor J, Wood K. Food intake and academic performance among adolescents. Can J Diet Pract Res. 2008;69(3):141–4. doi:10.3148/69.3.2008.141

39. López-Gil JF, Victoria-Montesinos D, García-Hermoso A. Is higher adherence to the mediterranean diet associated with greater academic performance in children and adolescents? A systematic review and meta-analysis. Clin Nutr. 2024;43(8):1702–9. doi:10.1016/j.clnu.2024.05.045

40. Martin A, Booth JN, Laird Y, Sproule J, Reilly JJ, Saunders DH. Physical activity, diet and other behavioural interventions for improving cognition and school achievement in children and adolescents with obesity or overweight. Cochrane Database of Systematic Reviews. 2018;2018(3). doi:10.1002/14651858.CD009728.pub4

41. McDiarmid K, Hodder R, Wolfenden L, Nathan N. The co-benefits of interventions to prevent obesity in children aged 6-18 years: A secondary data analysis. PROSPERO 2021 CRD42021281106 2021. https://www.crd.york.ac.uk/prospero/display_record.php?ID=CRD42021281106.

42. Harris PA, Taylor R, Thielke R, Payne J, Gonzalez N, Conde JG. Research electronic data capture (REDCap)—A metadata-driven methodology and workflow process for providing translational research informatics support. Journal of Biomedical Informatics. 2009;42(2):377–81. 10.1016/j.jbi.2008.08.010

43. Higgins JP, Altman DG. Assessing risk of bias in included studies. Cochrane handbook for systematic reviews of interventions: Cochrane book series. 2008:187–241.

44. Higgins JP, Eldridge S, Li T. Including variants on randomized trials. Cochrane handbook for systematic reviews of interventions. 2019:569–93.

45. Viechtbauer W. Conducting meta-analyses in R with the metafor package. Journal of statistical software. 2010;36:1–48.

46. Higgins JP, Li T, Deeks JJ. Choosing effect measures and computing estimates of effect. Cochrane handbook for systematic reviews of interventions. 2019:143–76.

47. Cohen J. Statistical Power Analysis for the Behavioural Sciences. Second ed. Hillsdale, N.J: L. Eribaum Associates; 1988.

48. Clemes SA, Bingham DD, Pearson N, et al. Public Health Research. Sit–stand desks to reduce sedentary behaviour in 9-to 10-year-olds: the Stand Out in Class pilot cluster RCT. Southampton (UK): NIHR Journals Library; 2020.

49. Neumark-Sztainer DRF, S. E. Flattum, C. F. Hannan, P. J. Story, M. T. Bauer, K. W. Feldman, S. B. Petrich, C. A. New moves-preventing weight-related problems in adolescent girls a group-randomized study. Am J Prev Med. 2010;39(5):421–32.

50. Neumark-Sztainer DS, M. Hannan, P. J. Rex, J. New Moves: a school-based obesity prevention program for adolescent girls. Prev Med. 2003;37(1):41–51.

51. Melnyk BMJ, D. Kelly, S. Belyea, M. Shaibi, G. Small, L. O’Haver, J. Marsiglia, F. F. Promoting healthy lifestyles in high school adolescents: a randomized controlled trial. Am J Prev Med. 2013;45(4):407–15.

52. Wilksch SMP, S. J. Byrne, S. M. Austin, S. B. McLean, S. A. Thompson, K. M. Dorairaj, K. Wade, T. D. Prevention Across the Spectrum: a randomized controlled trial of three programs to reduce risk factors for both eating disorders and obesity. Psychological Medicine. 2015;45(9):1811–23. doi: 10.1017/S003329171400289X

53. Donnelly JEG, J. L. Gibson, C. A. Smith, B. K. Washburn, R. A. Sullivani, D. K. et al. Physical Activity Across the Curriculum (PAAC): a randomized controlled trial to promote physical activity and diminish overweight and obesity in elementary school children. Prev Med 2009. p. 336–41.

54. Telford RDC, R. B. Fitzgerald, R. Olive, L. S. Prosser, L. Jiang, X. Telford, R. M. Physical education, obesity, and academic achievement: a 2-year longitudinal investigation of Australian elementary school children. Am J Public Health. 2012;102(2):368–74.

55. Gutin BY, Z. Johnson, M. Barbeau, P. Preliminary findings of the effect of a 3-year after-school physical activity intervention on fitness and body fat: the Medical College of Georgia Fitkid Project. International Journal of Pediatric Obesity 2008. p. 3–9.

56. Damsgaard CTD, S. M. Laursen, R. P. Ritz, C. Hjorth, M. F. Lauritzen, L. Sørensen, L. B. Petersen, R. A. Andersen, M. R. Stender, S. Andersen, R. Tetens, I. Mølgaard, C. Astrup, A. Michaelsen, K. F. Provision of healthy school meals does not affect the metabolic syndrome score in 8-11-year-old children, but reduces cardiometabolic risk markers despite increasing waist circumference. Br J Nutr. 2014;112(11):1826–36.

57. Muller I, Schindler C, Adams L, et al. Effect of a Multidimensional Physical Activity Intervention on Body Mass Index, Skinfolds and Fitness in South African Children: Results from a Cluster-Randomised Controlled Trial. Int J Environ Res Public Health. 2019;16(2):15. doi: 10.3390/ijerph16020232

58. Van Wijnen L, Wendel-Vos G, Wammes B, Bemelmans W. The impact of school-based prevention of overweight on psychosocial well-being of children. Obesity Reviews. 2009;10(3):298–312.

59. Andermo S, Hallgren M, Nguyen TT, et al. School-related physical activity interventions and mental health among children: a systematic review and meta-analysis. Sports Med Open. 2020;6(1):25. doi:10.1186/s40798-020-00254-x

60. Fu Q, Li L, Li Q, Wang J. The effects of physical activity on the mental health of typically developing children and adolescents: a systematic review and meta-analysis. BMC Public Health. 2025;25(1):1514. doi:10.1186/s12889-025-22690-8

61. García-Hermoso A, Ramírez-Vélez R, Lubans DR, Izquierdo M. Effects of physical education interventions on cognition and academic performance outcomes in children and adolescents: a systematic review and meta-analysis. Br J Sports Med. 2021;55(21):1224–32. doi:10.1136/bjsports-2021-104112

62. Li D, Wang D, Zou J, et al. Effect of physical activity interventions on children’s academic performance: a systematic review and meta-analysis. Eur J Pediatr. 2023;182(8):3587–601. doi:10.1007/s00431-023-05009-w

63. Watson A, Timperio A, Brown H, Best K, Hesketh KD. Effect of classroom-based physical activity interventions on academic and physical activity outcomes: a systematic review and meta-analysis. Int J Behav Nutr Phys Act. 2017;14(1):114. doi:10.1186/s12966-017-0569-9

