## Appendix I for "The effectiveness of school-based obesity prevention interventions on psychosocial and educative outcomes in children aged 6-18 years: a secondary data analysis"

**Appendix a**

**Table S1.** Characteristics of included studies

| Author year  Country | Study characteristics  Design (cluster type)  Setting  Age group years (mean age)  Gender | Number of participants (overall) | | Intervention  Treatment arms  Targeted behaviour  Theory  Duration | Comparator | Outcomes  Psychosocial or educative outcomes |
| --- | --- | --- | --- | --- | --- | --- |
|  |  | Randomised | Analysed |  |  |  |
| Adab 2018  United Kingdom | Design: C-RCT (School)  Setting: School * + Home + Community  Age group: 6-12 (6·3)  Gender: mixed | 2462 | 837 | Arms: 1  Target: DPA  Theory: not reported  Duration: ≤ 12 months | Control: usual practice | Psychosocial   - Quality of life - Body satisfaction |
| Bohnert 2013  USA | Design: RCT  Setting: School (ASP)  Age group: 6-12 (intv = 9·02, control = 9·38)  Gender: Girls only | 133 | 76 | Arms: 1  Target: DPA  Theory: SCT and Sociocultural theory  Duration: ≤ 12 months | Control: No intervention | Psychosocial   - Self-esteem - Body satisfaction |
| Breheny 2020  United Kingdom | Design: CRCT (School)  Setting: School  Age group: 6-12 years (8.9)  Gender: mixed | 2280 | 1670 | Arms: 1  Target: physical activity  Theory: BCT  Duration: ≤ 12 months | Control: No active intervention | Psychosocial   - Wellbeing - Quality of life   Educative   - Overall academic attainment |
| Clemes 2020  United Kingdom | Design: C-RCT (School)  Setting: School  Age group: 6-12 years (9·3)  Gender: mixed | 176 | 168 | Arms: 1  Target: physical activity  Theory: COM-B with BCW, TDF  Duration: ≤ 12 months | Control: Usual practice | Psychosocial   - Quality of life - Problem behaviours   Educative   - Maths - Reading - Writing - Engagement |
| Damsgaard 2014  Denmark | Design: C-RCT - crossover (School)  Setting: School  Age group: 6-12 (10·0)  Gender: mixed | 823 | 823 | Arms: 1  Target: Diet  Theory: not reported  Duration: ≤ 12 months | Control: Usual care (packed lunch from home) | Educative   - Maths - Reading - Attention and behaviour |
| De Greeff 2016  Netherlands | Design: CRCT (Classroom)  Setting: School  Age group: 6-12 years (8.1)  Gender: mixed | 376 | 376 | Arms: 1  Target: physical activity  Theory: not reported  Duration: ≤ 12 months | Control: Assume usual practice | Educative   - Maths - Reading |
| Dewar 2013  Australia | Design: C-RCT (School)  Setting: School* + Home  Age group: 13-18 (intv = 13·20, control = 13·15)  Gender: Girls only | 357 | 294 | Arms: 1  Target: DPA  Theory: SCT  Duration: ≤ 12 months | Control: Usual care presumed as no details but schoolbased intervention | Psychosocial   - Self-esteem |
| Donnelly 2009  USA | Design: C-RCT (School)  Setting: School  Age group: 6-12 (Grade 2: intv female = 7·7, control female = 7·8; intv male = 7·7, control male = 7·8. Grade 3: intv female = 8·7, control female = 8·7; intv male = 8·7, control male = 8·8)  Gender: mixed | 1527 | 1490 | Arms: 1  Target: physical activity  Theory: not reported  Duration: > 12 months | Control: Usual care - regular classroom instruction without  physically active lessons | Educative   - Maths - Reading - Writing |
| Dunker 2018  Brazil | Design: C-RCT (School)  Setting: School (ASP)* + Home  Age group: 13-18 (13·39)  Gender: Girls only | 270 | 270 | Arms: 1  Target: DPA  Theory: SCT  Duration: ≤ 12 months | Control: Usual practice | Psychosocial   - Self-esteem |
| Foster 2008  USA | Design: C-RCT (School)  Setting: School  Age group: 6-12 (intv = 11·13, control = 11·2)  Gender: mixed | 1349 | 843 | Arms: 1  Target: DPA  Theory: settings-based approach; CDC Guidelines to Promote Lifelong  Healthy Eating and physical activity  Duration: > 12 months | Control: No intervention | Psychosocial   - Body satisfaction |
| Gutin 2008  USA | Design: CRCT (School)  Setting: School  Age group: 6-12 years (8.5)  Gender: mixed | 601 | 447 | Arms: 1  Target: physical activity  Theory: Environmental change  Duration: > 12 months | Control: No intervention presumed as no details (afterschool  intervention) | Educative   - Maths - Reading - Writing |
| Harrington 2018  United Kingdom | Design: C-RCT (School)  Setting: School  Age group: 13-18 (12·8)  Gender: Girls only | 1753 | 1361 | Arms: 1  Target: physical activity  Theory: SCT  Duration: ≤ 12 months | Control: Usual practice | Psychosocial   - Quality of life - Self-esteem - Self-worth |
| Johnston 2013  USA | Design: CRCT (School)  Setting: School  Age group: 6-12 years (7-9 years, intervention = 7.8 ± 0.4, control = 7.7 ± 0.4)  Gender: mixed | 835 | 629 | Arms: 1  Target: DPA  Theory: not reported  Duration: > 12 months | Control: Self-help | Educative   - Overall academic attainment |
| Kennedy 2018  Australia | Design: C-RCT (School)  Setting: School* + Home  Age group: 13-18 (14·1)  Gender: mixed | 607 | 600 | Arms: 1  Target: physical activity  Theory: SCT and social-determination theory  Duration: ≤ 12 months | Control: Waitlist | Psychosocial   - Wellbeing - Self-esteem |
| Kriemler 2010  Switzerland | Design: C-RCT (School)  Setting: School* + Home  Age group: 6-12 (1st graders = 6·9; 5^th^ graders intv = 11·0, control = 11·3)  Gender: mixed | 502 | 502 | Arms: 1  Target: physical activity  Theory: SEM  Duration: ≤ 12 months | Control: Not informed of an intervention group | Psychosocial   - Quality of life |
| Kubik 2021  USA | Design: RCT  Setting: School (ASP)* + Home  Age group: 6-12 (9·3)  Gender: mixed | 132 | 122 | Arms: 1  Target: DPA  Theory: Social–ecological framework, healthy learner model for student chronic condition management  Duration: ≤ 12 months | Control: Newsletter only | Psychosocial   - Quality of life |
| Leme 2016  Brazil | Design: C-RCT (School)  Setting: School* + Home  Age group: 13-18 (16·05)  Gender: Girls only | 253 | 194 | Arms: 1  Target: DPA  Theory: SCT  Duration: ≤ 12 months | Control: Waitlist | Psychosocial   - Body satisfaction |
| Li 2019  China | Design: C-RCT (School)  Setting: School* + Home  Age group: 6-12 (intv = 6·15, control = 6·14)  Gender: mixed | 1641 | 1581 | Arms: 1  Target: DPA  Theory: Behaviour change techniques, social marketing principles, MRC framework  Duration: ≤ 12 months | Control: Usual practice | Psychosocial   - Quality of life |
| Lubans 2011  Australia | Design: C-RCT (School)  Setting: School* + Home  Age group: 13-18 (intv = 14·4, control = 14·2)  Gender: boys only | 100 | 100 | Arms: 1  Target: physical activity  Theory: SCT  Duration: ≤ 12 months | Control: Waitlist | Psychosocial   - Self-worth |
| Melnyk 2013  USA | Design: C-RCT (School)  Setting: School + Home  Age group: 13-18 (intv = 14·75, control = 14·74)  Gender: mixed | 807 | 627 | Arms: 1  Target: DPA  Theory: Cognitive theory  Duration: ≤ 12 months | Control: Attention control programme – safety and common health topics/issues | Psychosocial   - Anxiety - Depression |
| Muller 2019  South Africa | Design: C-RCT (School)  Setting: School  Age group: 6-12 (intv 1 = 10.0, intv 2 = 10.1, control = 9·9)  Gender: mixed | 1009 | 519 | Arms: 3  Target: physical activity  Theory: not reported  Duration: ≤ 12 months | Control: Usual practice | Psychosocial   - Wellbeing - Quality of life   Educative   - Overall academic attainment - Attention and behaviour |
| Neumark-Sztainer 2003  USA | Design: C-RCT (School)  Setting: School* + Home  Age group: 13-18 (intv = 14·9, control = 15·8)  Gender: Girls only | 201 | 190 | Arms: 1  Target: DPA  Theory: SCT  Duration: ≤ 12 months | Control: Regular physical education class and minimal intervention  (written materials on healthy eating  and physical activity at baseline) | Psychosocial   - Self-worth - Body satisfaction |
| Neumark-Sztainer 2010  USA | Design: C-RCT (School)  Setting: School* + Home  Age group: 13-18 (15·8)  Gender: Girls only | 356 | 336 | Arms: 1  Target: DPA  Theory: SCT, Stages of Change  Duration: > 12 months | Control: All-girls PE class during the first semester then usual PE | Psychosocial   - Self-worth - Body satisfaction |
| Sahota 2019  England | Design: C-RCT (School)  Setting: School  Age group: 6-12 (year 2 intv = 6·2, control = 6·3; year 4 = 8·3; overall 7·2)  Gender: mixed | 358 | 311 | Arms: 1  Target: DPA  Theory: Behaviour Theory, BCW  Duration: > 12 months | Control: Usual practice | Psychosocial   - Body satisfaction |
| Sallis 1993  USA | Design: C-RCT (School)  Setting: School  Age group: 6-12 (9·25)  Gender: mixed | 745 | 549 | Arms: 2  Target: physical activity  Theory: Behaviour Change and self-management  Duration: > 12 months | Control: Usual care PE | Educative   - Overall academic attainment - Maths - Reading |
| Smith 2014  Australia | Design: C-RCT (School)  Setting: School* + Home  Age group: 13-18 (12·7)  Gender: Boys only | 361 | 361 | Arms: 1  Target: physical activity  Theory: Self-determination theory and SCT  Duration: ≤ 12 months | Control: Waitlist and usual practice (i.e. regularly scheduled  school sports and PE) | Psychosocial   - Wellbeing |
| Story 2003a  USA | Design: RCT  Setting: School* + Home  Age group: 6-12 (intv = 9·4, control = 9·1)  Gender: Girls only | 53 | 53 | Arms: 1  Target: DPA  Theory: SCT, youth development, and resiliency based approach  Duration: ≤ 12 months | Control: “active placebo,” non-nutrition/physical activity condition, promoting self-esteem and cultural enot reportedichment | Psychosocial   - Body satisfaction |
| Telford 2012  Australia | Design: C-RCT (Schools)  Setting: School  Age group: 6-12 (not reported)  Gender: mixed | Unclear | 620 | Arms: 1  Target: physical activity  Theory: not reported  Duration: > 12 months | Control: Usual care, common practice PE | Educative   - Maths - Reading - Writing |
| Velez 2010  USA | Design: RCT  Setting: School  Age group: 13-18 years (intervention + control: 16.14 ± 0.19)  Gender: mixed | 31 | 28 | Arms: 1  Target: physical activity  Theory: not reported  Duration: ≤ 12 months | Control: No intervention | Psychosocial   - Self-worth |
| Waters 2017  Australia | Design: C-RCT (School)  Setting: School* + Community + Home  Age group: 6-12 (not reported)  Gender: mixed | 3222 | 2743 | Arms: 1  Target: DPA  Theory: Health Promoting Schools Framework (based on health promotion theory and consistent with a socio-environmental theoretical framework) and International Obesity Task Force ‘10 guiding principles for obesity prevention'  Duration: > 12 months | Control: Usual practice | Psychosocial   - Wellbeing - Quality of life |
| White 2019  USA | Design: RCT  Setting: Community* + Home  Age group: 6-12 (9·35)  Gender: mixed | 228 | 125 | Arms: 1  Target: DPA  Theory: SCT, experiential 4-H learning model  Duration: > 12 months | Control: No intervention | Psychosocial   - Quality of life |
| Wilksch 2015  Australia | Design: C-RCT (Classroom)  Setting: School  Age group: 13-18 (13·21)  Gender: mixed | 820 | 820 | Arms: 1  Target: DPA  Theory: not reported  Duration: ≤ 12 months | Control: Usual school class | Psychosocial   - Depression - Body satisfaction |

*Majority setting

ASE = Attitude, social influence and self-efficacy model, ASP = after school program, BCT = Behaviour Change Theories, BCW = Behaviour Change Wheel, CDC = Centres for Disease Control, COM-B = capability, opportunity, motivation – behaviour, CMT = competence motivation theory, C-RCT = cluster randomised controlled trial, DPA = diet and physical activity, HPSF = Health Promoting Schools Framework, IMB model = information-motivation-behavioural skills model, Intv = intervention, MRC = medical research council, RCT = randomised controlled trial, SEM = Social Ecological Model, SCT = Social Cognitive Theory, SLT = Social learning theory, TDF = theoretical domains framework, TPB = Theory of planned behaviour, TTM = Transtheoretical model (Stages of Change), USA = United States of America
