## Appendix II for "The effectiveness of school-based obesity prevention interventions on psychosocial and educative outcomes in children aged 6-18 years: a secondary data analysis"

**Appendix b**

**Table S2.** Risk of bias of included studies assessing school obesity prevention interventions in children from 1990-2023

| Study Identification | Design | Intervention type | Outcome | Random sequence (selection bias) | Allocation concealment (selection bias) | Blinding (performance and detection bias) | Incomplete outcome data (attrition bias) | Selective outcome reporting  (reporting bias) | Other overall |
| --- | --- | --- | --- | --- | --- | --- | --- | --- | --- |
| Adab 2018 | C-RCT | DPA | Quality of life | Low | Low | High | Low | Low | High |
|  |  |  | Body satisfaction | Low | Low | High | Low | Low | High |
| Bohnert 2013 | RCT | DPA | Self-esteem | High | High | High | High | Unclear | High |
|  |  |  | Body satisfaction | High | High | High | High | Unclear | High |
| Breheny 2020 | C-RCT | PA | Wellbeing | Low | Unclear | High | High | Low | High |
|  |  |  | Quality of life | Low | Unclear | High | High | Low | High |
| Clemes 2020 | C-RCT | PA | Quality of life | Low | Unclear | High | Low | Low | High |
|  |  |  | Problem behaviours | Low | Unclear | High | Low | Low | High |
|  |  |  | Maths | Low | Unclear | High | Low | Low | High |
|  |  |  | Reading | Low | Unclear | High | Low | Low | High |
|  |  |  | Writing | Low | Unclear | High | Low | Low | High |
|  |  |  | Engagement | Low | Unclear | High | Low | Low | High |
| Damsgaard 2014 | C-RCT | D | Attention and behaviour | Low | High | High | Unclear | Low | High |
|  |  |  | Maths | Low | High | High | Unclear | Low | High |
|  |  |  | Reading | Low | High | High | Unclear | Low | High |
| De Greeff 2016 | C-RCT | PA | Maths | Unclear | Low | Unclear | Low | Unclear | Unclear |
|  |  |  | Reading | Unclear | Low | Unclear | Low | Unclear | Unclear |
| Dewar 2013 | C-RCT | DPA | Self-esteem | Unclear | Low | High | Low | Low | High |
| Donnelly 2009 | C-RCT | PA | Maths | Unclear | Unclear | Unclear | Unclear | Unclear | Unclear |
|  |  |  | Reading | Unclear | Unclear | Unclear | Unclear | Unclear | Unclear |
|  |  |  | Writing | Unclear | Unclear | Unclear | Unclear | Unclear | Unclear |
| Dunker 2018 | C-RCT | DPA | Self-esteem | Unclear | Unclear | High | Low | Low | High |
| Foster 2008 | C-RCT | DPA | Body satisfaction | Unclear | Unclear | High | High | Unclear | High |
| Gutin 2008 | C-RCT | PA | Maths | Low | High | Unclear | Low | High | High |
|  |  |  | Reading | Low | High | Unclear | Low | High | High |
|  |  |  | Writing | Low | High | Unclear | Low | High | High |
| Harrington 2018 | C-RCT | PA | Quality of life | Low | Low | High | High | Low | High |
|  |  |  | Self-esteem | Low | Low | High | High | Low | High |
|  |  |  | Self-worth | Low | Low | High | High | Low | High |
| Johnston 2013 | C-RCT | DPA | Overall academic achievement | Low | Unclear | High | High | Unclear | High |
| Kennedy 2018 | C-RCT | PA | Wellbeing | Low | Unclear | High | Low | Low | High |
|  |  |  | Self-esteem | Low | Unclear | High | Low | Low | High |
| Kriemler 2010 | C-RCT | PA | Quality of life | Low | Low | High | Low | Low | High |
| Kubik 2021 | RCT | DPA | Quality of life | Low | Unclear | High | Low | Low | High |
| Leme 2016 | C-RCT | DPA | Body satisfaction | Low | Low | High | High | Low | High |
| Li 2019 | C-RCT | DPA | Quality of life | Low | Unclear | High | Low | Low | High |
| Lubans 2011 | C-RCT | PA | Self-worth | Unclear | Unclear | High | Low | Low | High |
| Melnyk 2013 | C-RCT | DPA | Anxiety | Unclear | Unclear | Unclear | Low | Low | Unclear |
|  |  |  | Depression | Unclear | Unclear | Unclear | Low | Low | Unclear |
| Muller 2019 | C-RCT | PA | Attention and behaviour | Low | Unclear | Unclear | High | Low | High |
|  |  |  | Overall academic achievement | Low | Unclear | Unclear | High | Low | High |
|  |  |  | Quality of life | Low | Unclear | High | High | Low | High |
|  |  |  | Wellbeing | Low | Unclear | High | High | Low | High |
| Neumark-Sztainer 2003 | C-RCT | DPA | Body satisfaction | Unclear | High | High | Low | Unclear | High |
|  |  |  | Self-worth | Unclear | High | High | Low | Unclear | High |
| Neumark-Sztainer 2010 | C-RCT | DPA | Body satisfaction | Unclear | Unclear | High | Low | Low | High |
|  |  |  | Self-worth | Unclear | Unclear | High | Low | High | High |
| Sahota 2019 | C-RCT | DPA | Body satisfaction | Low | Unclear | High | Low | Unclear | High |
| Sallis 1993 | C-RCT | PA | Maths | High | Unclear | Unclear | High | Unclear | High |
|  |  |  | Overall academic achievement | High | Unclear | Unclear | High | Unclear | High |
|  |  |  | Reading | High | Unclear | Unclear | High | Unclear | High |
| Smith 2014 | C-RCT | PA | Wellbeing | Low | Low | High | High | Low | High |
| Story 2003a | RCT | DPA | Body satisfaction | Low | Unclear | High | High | Low | High |
| Telford 2012 | C-RCT | PA | Maths | Unclear | Unclear | Unclear | Unclear | Low | Unclear |
|  |  |  | Reading | Unclear | Unclear | Unclear | Unclear | Low | Unclear |
|  |  |  | Writing | Unclear | Unclear | Unclear | Unclear | Low | Unclear |
| Velez 2010 | RCT | PA | Self-worth | Unclear | Unclear | Unclear | Unclear | Unclear | Unclear |
| Waters 2017 | C-RCT | DPA | Quality of life | Low | Low | High | Low | Low | High |
|  |  |  | Wellbeing | Low | Low | High | Low | Low | High |
| White 2019 | RCT | DPA | Quality of life | Low | Unclear | High | high | Unclear | High |
| Wilksch 2015 | C-RCT | DPA | Body satisfaction | Unclear | Unclear | High | High | Unclear | High |
|  |  |  | Depression | Unclear | Unclear | High | High | Unclear | High |

RCT = randomised controlled trial; C-RCT = cluster-randomised controlled trial; D = diet; PA = physical activity; DPA = diet and physical activity
